# Associations between Psychological Factors and Long-Term Dizziness Handicap in Vestibular Schwannoma Patients: A Cross-Sectional Study

**DOI:** 10.1101/2024.10.19.24315803

**Authors:** M. Rutenkröger, M. Scheer, S. Rampp, C. Strauss, R. Schönfeld, B. Leplow

**Affiliations:** Department of Medical Psychology, University Medical Center Hamburg-Eppendorf, Martinistraße 52, 20246 Hamburg; Department of Neurosurgery, University Hospital Halle, Ernst-Grube-Straße 40, 06120 Halle; Department of Neurosurgery, University Hospital Erlangen, Schwabachanlage 6, 91054 Erlangen; Department of Neuroradiology, University Hospital Erlangen, Schwabachanlage 6, 91054 Erlangen, Germany; Department of Psychology, Martin-Luther-Universität Halle-Wittenberg, Emil-Abderhalden-Straße 26-27, 06108 Halle

**Author notes:** Corresponding Author’s name and current institution: Mareike Rutenkröger, Department of Medical Psychology, University Medical Center Hamburg-Eppendorf, Martinistraße 52, 20246 Hamburg, Corresponding Author’s.

**Keywords:** vestibular schwannoma, dizziness, vertigo, microsurgery, personality, psychological factors

## Abstract

**Background:** Vestibular schwannoma (VS) often results in persistent dizziness that negatively impacts quality of life (QoL). While physical effects are well-documented, the influence of psychological factors on dizziness severity is less studied. This study investigates how psychological characteristics affect dizziness in VS patients.

**Methods:** In this cross-sectional study, 93 VS patients were analyzed, with 77 (82.8%) reporting postoperative dizziness. Psychological factors, including premorbid disorders, personality traits, somatization, and current depression and anxiety levels, were assessed using self-report questionnaires. Correlations between these factors and dizziness severity, measured by the Dizziness Handicap Inventory (DHI), were examined.

**Results:** Patients with postoperative dizziness had higher depression levels and a greater prevalence of preoperative dizziness compared to those without postoperative dizziness. Significant correlations were found between dizziness severity and psychological factors: conscientiousness (r = .30, p = .03), social support (r = .32, p = .03), and the HADS total score (r = .36, p = .01). Emotional aspects of dizziness (DHI-E) were correlated with somatization (r = .27, p = .04) and anxiety (r = .40, p = .01). Functional aspects (DHI-F) were linked to conscientiousness (r = .31, p = .03) and depression (r = .26, p = .06).

**Conclusion:** Psychological factors significantly impact dizziness severity in VS patients. Incorporating psychological assessments and interventions, such as cognitive-behavioral therapy and combined vestibular and psychological rehabilitation, may improve treatment outcomes and QoL. Further research is needed to assess the effectiveness of these approaches and their impact on the relationship between psychological factors and dizziness.

## Introduction

A vestibular schwannoma (VS), also referred to as an acoustic neuroma, is a benign neoplasm that arises from the Schwann cells of the vestibulocochlear nerve. While the primary consequence of this condition is often auditory, many patients also present with dizziness and vertigo, which may persist after or be induced by microsurgery or radiation treatment (1–3). Conservative VS management (“wait-and-scan”) seems to have no impact on the prevalence of these symptoms, as well as postural sway and canal paresis (4). The severity of these vestibular symptoms is variable, but they frequently have a long-lasting impact on patients’ quality of life (QoL) across multiple domains (2,3,5).

As identified by Carlson et al. (5), the strongest predictors of long-term QoL reduction in patients with sporadic VS are ongoing dizziness and headache. In comparison, the impact of hearing loss, facial nerve function, and tinnitus was shown to be less significant (5). Dizziness is significantly correlated with challenges in performing routine tasks, which can lead to a deterioration in physical well-being, an increased reliance on external assistance, and occupational limitations (6). It has been demonstrated that dizziness is linked to elevated levels of anxiety, depression, and diminished cognitive function, which serves to further exacerbate the decline in QoL (6,7). Furthermore, balance disorders were found to be associated with anxiety and depression, as well as with greater daily consequences and a denial coping response (8). In an experimental setting, the presence of anxiety symptoms during a vestibular stimulus may contribute to a priming effect that could explain the observed worsening of balance function (9).

In addition to the dizziness associated with VS, it has also been shown that somatoform complaints in individuals experiencing dizziness are highly prevalent. The presence of certain factors, such as personality characteristics or accompanying psychopathology, has been identified as influencing the prevalence of these complaints (10). Research on chronic subjective dizziness (11) and persistent postural-perceptual dizziness (PPPD) (12) has revealed that anxiety, neuroticism, and somatic symptom burden are markedly elevated in individuals with these conditions. Patients with PPPD also exhibited diminished levels of conscientiousness in comparison to the control group (12). In the aftermath of acute vestibular events, psychological and behavioral responses, along with brain maladaptation, were shown to be the most probable predictors of PPPD (13). Furthermore, a correlation was identified between benign paroxysmal positional vertigo and the presence of comorbid major depressive disorder, generalized anxiety disorder, and obsessive-compulsive personality disorders when compared to the findings in a control group (14).

Given the lack of comprehensive examination of psychological factors in this area, the objective of this study was to evaluate the differential impact of these factors on dizziness and to ascertain the potential interplay between psychological characteristics and the severity of dizziness handicap. Specifically, the present study aimed to: 1) elucidate the distinctions between VS patients with and without postoperative dizziness. Accordingly, our investigation concentrated on the incidence of premorbid psychological disorders and ailments, personality traits, somatization tendencies, and the current levels of depression and anxiety. And, 2) examine the relationships between the intensity of dizziness handicap and these psychological variables as well as sociodemographic variables (age, gender) and tumor size within the group of individuals affected by dizziness.

## Materials and Methods

This cross-sectional, single-center investigation was conducted at University Hospital Halle (Saale), Germany, as part of a broader study that also explored psychological factors and their associations with postoperative headache and tinnitus in patients with VS patients (15). The inclusion criteria were as follows: individuals with a confirmed diagnosis of vestibular schwannoma (VS), aged 18 or above at the time of diagnosis, undergoing retrosigmoid surgery, and possessing proficiency in the German language. Patients were excluded if they had undergone prior surgical procedures or radiation therapy, if they had a history of recurrent VS, if they had been diagnosed with additional oncological conditions, or if they had been diagnosed with neurofibromatosis type 2. The advent of the SARS-CoV-2 pandemic precipitated a postponement in the recruitment of subjects, with recruitment commencing on 01/05/2020, and concluding on 31/08/2020. The initial stage of participant engagement was conducted through direct approaches during resident consultations at University Hospital Halle, and an online survey (*SoSciSurvey*) comprising identical questions was administered by *Vereinigung Akustikus Neurinom e*.*V*. (a non-profit patient self-help organization). Prior to participation, all subjects provided written informed consent. The study was approved by the Ethics Committee of University Hospital Halle (No. 2020-008).

### Measures

A self-administered survey was employed to collect data pertaining to demographic characteristics (e.g., age and gender), psychological aspects (including premorbid psychological diagnoses such as anxiety disorders or other mental conditions such as sleep disturbances), and treatment details (e.g., surgical approach, craniotomy vs. craniectomy). The survey consisted of binary questions (yes/no), with the option of providing additional details when necessary (e.g., “Specify the diagnosed psychological disorder”). Tumor size was evaluated according to the Koos grading system, which ranges from 1 to 4 (16).

The German version of the Dizziness Handicap Inventory, known as DHI-G (17), was employed for the purpose of evaluating the extent of disability associated with dizziness. The scale comprises 25 items, with a score of 4 allocated to affirmative responses, 2 to “sometimes” responses, and 0 to negative responses. The total score ranges from 0, indicating no disability, to 100, reflecting severe disability. The questionnaire is comprised of three subscales: a seven-item physical subscale, a nine-item emotional subscale, and a nine-item functional subscale. The DHI-G has been demonstrated to perform reliably, and it is therefore recommended as a suitable assessment tool for gauging disability in individuals experiencing dizziness and unsteadiness.

The Glasgow Benefit Inventory (GBI) is a validated questionnaire comprising 18 items that is utilized to assess patients’ perceived health benefits (PHB) post-intervention, such as surgery. GBI scores range from -100 to +100, with a score of 0 indicating no perceived benefit, +100 signifying the highest level of benefit, and negative scores suggesting a decline in health. Principal component analysis revealed that the Glasgow Benefit Inventory items consistently fell into three distinct subscales. The first subscale, labeled “General,” comprised twelve questions related to overall health changes, both general and psychosocial. The second subscale, labeled “Social,” included three questions addressing the necessary social support for the specific condition at hand. The final subscale, labeled “Physical,” included three questions focusing on changes in physical health status, including medication requirements and doctor visits (18).

In order to assess the Big Five personality traits in accordance with the framework established by McCrae and Costa (19), the German version of the Ten Item Personality Inventory (TIPI-G) (20) was utilized. This inventory captures the five fundamental dimensions of the personality model, namely emotional stability (the opposite of neuroticism), extraversion, agreeableness, openness, and conscientiousness. The TIPI-G, which was designed as a concise version of the questionnaire, is particularly beneficial in situations with time constraints, as it provides a reliable estimate of the comprehensive measurements associated with the five-factor model of personality, such as those provided by the NEO-PI-R (21). The respondents employ a 7-point Likert scale (ranging from 1 for “strongly disagree” to 7 for “strongly agree”) to evaluate ten pairs of adjectives. The resulting scale score is the mean of two items, including one negatively worded item.

Pre-morbid somatization tendencies were assessed using the Screening for Somatoform Disorders (SOMS-2), a validated instrument commonly used in patient populations with psychosomatic disorders (22). In the first part of the questionnaire, participants were asked to report physical symptoms that were intermittent or persistent in the two years prior to the diagnosis of VS. These symptoms should have significantly interfered with their well-being or personal lifestyle, without a clearly identified cause as determined by medical professionals. The questionnaire presented a list of 53 somatoform symptoms, 5 specific to women and 1 specific to men, based on criteria from the Diagnostic and Statistical Manual of Mental Disorders (DSM-IV-TR) and the International Classification of Diseases (ICD-10). The second section consisted of 15 questions designed to assess disability, frequency of medical consultations due to symptoms, and inclusion/exclusion criteria for all somatoform disorders.

We used the Hospital Anxiety and Depression Scale (HADS-D) to assess current symptoms of anxiety and depression (23). This questionnaire consists of two scales, one for anxiety and the other for depression, each ranging from 0 to 21. A total score, ranging from 0 to 42, was calculated based on symptoms experienced in the week prior to the assessment. Elevated scores indicated increased levels of anxiety, depression, or general psychological distress. The HADS-D serves as a screening tool, with scores greater than 8 warranting additional assessment for an affective disorder.

The statistical analysis was conducted using the statistical software package SPSS (24), with a significance level set at α = 0.05. The normality of the dataset was evaluated through the application of the Shapiro-Wilk test, which indicated a non-normal distribution. As the objective of this explorative study was to gain a more comprehensive understanding of the relationships and differences among psychological factors and postoperative dizziness, we combined Chi-square tests, Mann-Whitney U-tests and correlation analyses. Chi-square tests were employed to ascertain discrepancies between the dizziness and non-dizziness groups with respect to dichotomous variables, including the presence of premorbid psychological disorders. Mann-Whitney U-tests provide insights into the differences between patient groups with and without dizziness, allowing us to identify significant disparities. In contrast, the use of Spearman correlation analysis (r_s_) within the dizziness group facilitates a more comprehensive examination of the precise dynamic relationships between these variables, thereby providing insight into how they interact and potentially influence the severity or nature of dizziness. This dual approach enables a more nuanced interpretation of our findings, capturing both intergroup differences and intragroup associations. To address multiple testing, false discovery rate correction (25) was applied.

## Results

### Sample characteristics

A total of 93 participants were included in the final analysis, following the exclusion of eight individuals whose questionnaires were incomplete or lacked data regarding tumor size. Of the evaluated participants, 77 (82.8%) reported experiencing postoperative dizziness. It is noteworthy that over half of the patients surveyed indicated that they had also experienced dizziness prior to surgery. The participants’ ages ranged from 23 to 85 years, with a mean age of 55.4 years (SD = 11.9). The mean age of the participants at the time of surgery was 46.7 years (SD = 10.2), and the mean duration since treatment was 7.6 years (SD = 7.8). The preoperative dizziness severity scores, as measured by the Numerical Analog Scale (NAS), ranged from 1 to 10, with a mean score of 4.6 (SD = 2.7). Similarly, postoperative dizziness severity scores ranged from 1 to 10, with a mean score of 4.8 (SD = 2.7). A total of 18.2% of participants reported experiencing a severe degree of dizziness-related handicap. All patients who reported preoperative dizziness also experienced postoperative dizziness, and 29 patients (37.6%) reported that dizziness began to occur after surgery. Table 1 presents the descriptive statistics for the additional demographic variables and questionnaire scales. The descriptive data for the administered questionnaires are presented in Table 2.

**Table 1.**
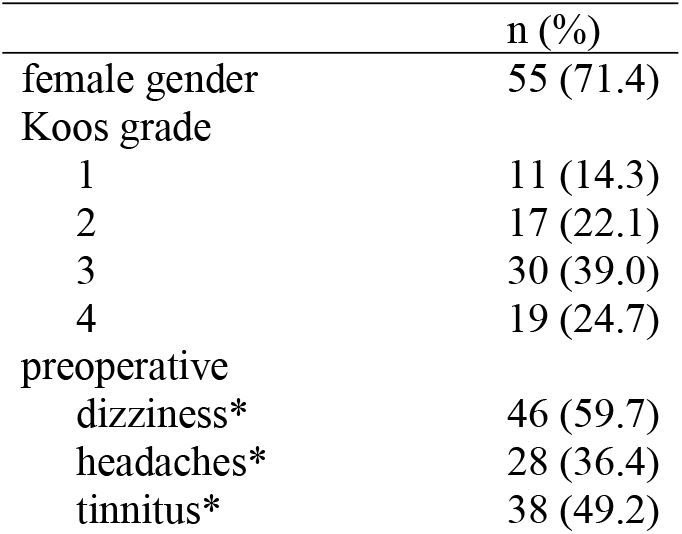
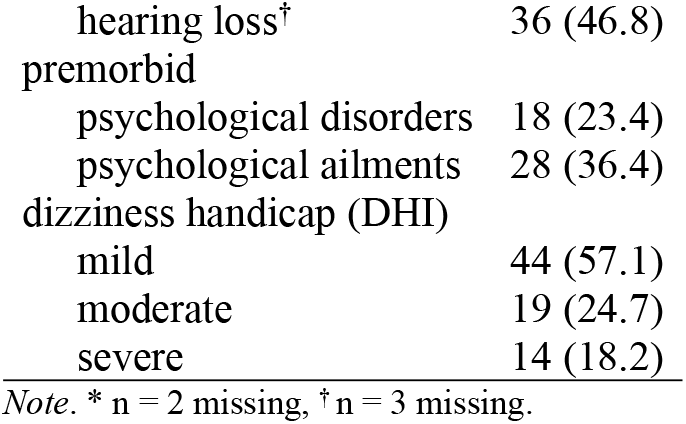
Absolute and relative frequencies of demographic and dizziness handicap-related variables (*N*=77).

**Table 2.** Descriptive data used questionnaires (*N* = 77)

| Questionnaire scales | Minimum | Maximum | M (SD) |
| --- | --- | --- | --- |
| *DHI-G* |  |  |  |
| physical | 0 | 28 | 11.3 (6.9) |
| emotional | 0 | 28 | 9.4 (7.4) |
| functional | 0 | 36 | 12.4 (8.2) |
| total score | 1 | 92 | 33.1 (20.6) |
| *TIPI-G* |  |  |  |
| extraversion | 2 | 14 | 8.2 (3.1) |
| agreeableness | 6 | 14 | 10.8 (2.1) |
| conscientiousness | 7 | 14 | 12.4 (1.6) |
| emotional stability | 2 | 14 | 9.9 (3.0) |
| openness | 4 | 14 | 10.3 (2.7) |
| *GBI* |  |  |  |
| global | -100 | 75 | -19.8 (33.8) |
| social support | -83 | 67 | 1.7 (26.2) |
| physical health | -67 | 33 | -16.5 (28.6) |
| overall | -81 | 64 | -15.7 (24.8) |
| *SOMS-2* |  |  |  |
| symptom count score | 1 | 35 | 9.4 (7.3) |
| somatization index ICD-10 | 1 | 14 | 3.8 (3.3) |
| somatization index DSM-IV | 1 | 22 | 3.3 (4.2) |
| *HADS-D* |  |  |  |
| depression | 0 | 21 | 4.9 (3.5) |
| anxiety | 0 | 18 | 7.3 (3.7) |
| total score | 3 | 39 | 12.2 (6.4) |

Abbreviations. DHI-G = Dizziness Handicap Inventory; TIPI-G = Ten Item Personality Inventory-German; GBI = Glasgow Benefit Inventory; SOMS-2 = Screening for Somatoform Disorders; HADS-D = Hospital Anxiety and Depression Scale.

### Differences between patients with and without postoperative dizziness

A comparison of patients with and without postoperative dizziness revealed that those experiencing dizziness following surgery exhibited a higher mean symptom count score (M = 9.4, SD = 7.3 vs. M = 5.4, SD = 4.7, p < .05) and higher levels of depression (M = 7.3, SD = 3.8 vs. M = 5.1, SD = 3.8, p < .05). No significant differences were identified with regard to personality traits or perceived health benefits. Patients who experienced postoperative dizziness exhibited a significantly higher prevalence of preoperative dizziness (n = 46 vs. 2, p = .001) and preoperative headaches (n = 28 vs. 1, p < .05) compared to those who did not.

### Associations between postoperative dizziness handicap and psychological factors

After correcting for multiple comparisons, the analysis yielded several noteworthy correlations (see Table 3). With regard to the physical aspects of dizziness (DHI-P), the following significant correlations were identified: Conscientiousness demonstrated a positive correlation (r = .30, corrected p = .03), as did social support (r = .32, corrected p = .02). With regard to the emotional aspects of dizziness (DHI-E), the following significant correlations were identified: The Somatization Index DSM-IV demonstrated a correlation of r = .27 (corrected p = .04), while the HADS Depression and HADS Anxiety exhibited correlations of r = .31 (corrected p = .03) and r = .40 (corrected p = .01), respectively. For the functional aspects of dizziness (DHI-F), significant correlations were observed with conscientiousness (r = .31, corrected p = .03), and the somatization index DSM-IV (r = .30, corrected p = .03).The overall dizziness handicap (DHI total score) demonstrated significant correlations with conscientiousness (r = .30, corrected p = .03), social support (r = .32, corrected p = .03), and the HADS total score (r = .36, corrected p = .01).

**Table 3.**
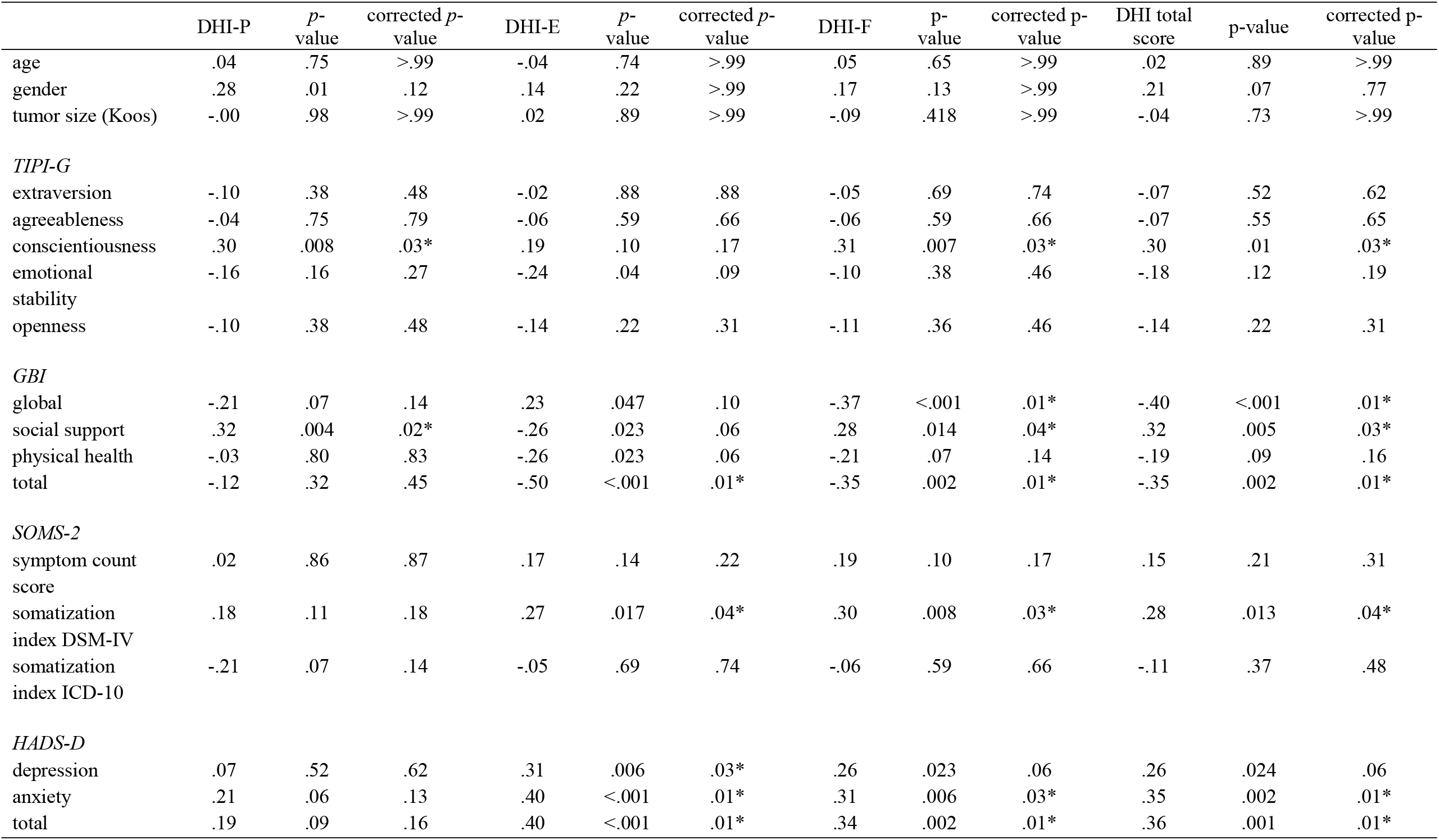
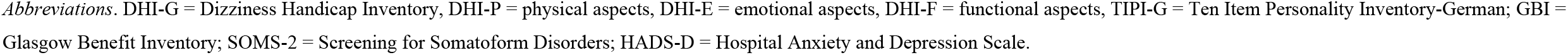
Correlation matrix for dizziness handicap (DHI-G), personality (TIPI-G), perceived health benefit (GBI), somatization tendencies (SOMS-2) and psychological burden (HADS-D)

|  | DHI-P | *p*-value | corrected *p*-value | DHI-E | *p*-value | corrected *p*-value | DHI-F | *p*-value | corrected *p*-value | DHI total score | *p*-value | corrected *p*-value |
| --- | --- | --- | --- | --- | --- | --- | --- | --- | --- | --- | --- | --- |
| age | .04 | .75 | >.99 | -.04 | .74 | >.99 | .05 | .65 | >.99 | .02 | .89 | >.99 |
| gender | .28 | .01 | .12 | .14 | .22 | >.99 | .17 | .13 | >.99 | .21 | .07 | .77 |
| tumor size (Koos) | -.00 | .98 | >.99 | .02 | .89 | >.99 | -.09 | .418 | >.99 | -.04 | .73 | >.99 |
| *TIPI-G* |  |  |  |  |  |  |  |  |  |  |  |  |
| extraversion | -.10 | .38 | .48 | -.02 | .88 | .88 | -.05 | .69 | .74 | -.07 | .52 | .62 |
| agreeableness | -.04 | .75 | .79 | -.06 | .59 | .66 | -.06 | .59 | .66 | -.07 | .55 | .65 |
| conscientiousness | .30 | .008 | .03* | .19 | .10 | .17 | .31 | .007 | .03* | .30 | .01 | .03* |
| emotional stability | -.16 | .16 | .27 | -.24 | .04 | .09 | -.10 | .38 | .46 | -.18 | .12 | .19 |
| openness | -.10 | .38 | .48 | -.14 | .22 | .31 | -.11 | .36 | .46 | -.14 | .22 | .31 |
| *GBI* |  |  |  |  |  |  |  |  |  |  |  |  |
| global | -.21 | .07 | .14 | .23 | .047 | .10 | -.37 | <.001 | .01* | -.40 | <.001 | .01* |
| social support | .32 | .004 | .02* | -.26 | .023 | .06 | .28 | .014 | .04* | .32 | .005 | .03* |
| physical health | -.03 | .80 | .83 | -.26 | .023 | .06 | -.21 | .07 | .14 | -.19 | .09 | .16 |
| total | -.12 | .32 | .45 | -.50 | <.001 | .01* | -.35 | .002 | .01* | -.35 | .002 | .01* |
| *SOMS-2* |  |  |  |  |  |  |  |  |  |  |  |  |
| symptom count score | .02 | .86 | .87 | .17 | .14 | .22 | .19 | .10 | .17 | .15 | .21 | .31 |
| somatization index DSM-IV | .18 | .11 | .18 | .27 | .017 | .04* | .30 | .008 | .03* | .28 | .013 | .04* |
| somatization index ICD-10 | -.21 | .07 | .14 | -.05 | .69 | .74 | -.06 | .59 | .66 | -.11 | .37 | .48 |
| *HADS-D* |  |  |  |  |  |  |  |  |  |  |  |  |
| depression | .07 | .52 | .62 | .31 | .006 | .03* | .26 | .023 | .06 | .26 | .024 | .06 |
| anxiety | .21 | .06 | .13 | .40 | <.001 | .01* | .31 | .006 | .03* | .35 | .002 | .01* |
| total | .19 | .09 | .16 | .40 | <.001 | .01* | .34 | .002 | .01* | .36 | .001 | .01* |

*Abbreviations.* DHI-G = Dizziness Handicap Inventory, DHI-P = physical aspects, DHI-E = emotional aspects, DHI-F = functional aspects, TIPI-G = Ten Item Personality Inventory-German; GBI = Glasgow Benefit Inventory; SOMS-2 = Screening for Somatoform Disorders; HADS-D = Hospital Anxiety and Depression Scale.

## Discussion

To the best of our knowledge, this is the first study to examine the associations between long-term dizziness and psychological factors, such as personality and somatization, in VS patients who underwent retrosigmoid microsurgery. An understanding of these relationships is essential for the development of more effective treatment strategies that address both the physical and psychological dimensions of dizziness in this patient population.

The observation that a significant proportion of patients experienced dizziness both pre- and postoperatively is consistent with previous research indicating that pre-existing vestibular symptoms are a strong predictor of postoperative dizziness (1–3). The elevated mean symptom count score and levels of depression in patients with postoperative dizziness serve to underscore the lasting impact of these symptoms on patients’ well-being. This finding supports the assertion of Carlson et al. that persistent dizziness is a critical predictor of long-term reduction in quality of life in VS patients (5). However, in contrast to the findings of Carlson et al, we found no significant associations between age, gender, tumor size, and dizziness handicap.

The significant correlations between dizziness and psychological factors offer a more profound comprehension of dizziness in VS patients. Specifically, a positive correlation was observed between conscientiousness and the physical and functional aspects of dizziness of patients with VS. This indicates that patients with higher levels of conscientiousness may experience disparate patterns of dizziness or demonstrate more effective coping mechanisms. In contrast, elevated levels of depression and anxiety were markedly linked to high levels of both emotional and functional dizziness handicap. These findings are consistent with those of previous studies that have highlighted the role of psychological distress in exacerbating dizziness and its associated disability (6–8).

The correlation between the somatization index (DSM-IV) and the emotional aspects of dizziness provides further evidence that somatic symptoms and psychological distress might be closely linked. This is consistent with findings from research on chronic subjective dizziness and PPPD, which have demonstrated elevated levels of somatic symptom burden in these conditions (11, 12).

The positive correlation between social support and the physical and overall handicap of dizziness indicates that patients with more robust social support networks may experience less severe physical symptoms and overall handicap. This finding is consistent with the results of previous research indicating that social support can mitigate the impact of chronic conditions on quality of life (7).

The significant correlations identified in this study underscore the necessity of addressing psychological factors in the management of dizziness in VS patients. The integration of psychological assessments and interventions, such as cognitive-behavioral therapy or vestibular rehabilitation combined with psychological support, may prove beneficial especially in those with premorbid dizziness problems and/or other psychophysiological/emotional problems. This approach could assist in addressing the multifaceted nature of dizziness, thereby improving both physical symptoms and overall QoL. In accordance with the recommended treatment strategies for PPPD, psychotherapy should incorporate an educational component that elucidates the manner in which symptoms arise from altered sensory processing and postural control, frequently precipitated by a past stressor or vestibular disorder. Although maladaptive beliefs and postural habits may have developed, they are susceptible to modification. While complete remission of symptoms may not always be attainable, a substantial improvement remains a realistic and promising outcome (26).

### Strength and limitations

This study’s principal strength is its comprehensive investigation of a range of psychological variables, including premorbid psychological disorders, personality traits, somatization tendencies, and current levels of depression and anxiety. By integrating these factors, the study provides a nuanced understanding of how psychological characteristics interact with dizziness severity in VS patients. The study identified several significant correlations between psychological factors and different aspects of dizziness, which adds valuable insight into the complex relationship between these variables. For instance, the observed associations between conscientiousness, social support, and the severity of dizziness highlight the multifaceted nature of dizziness and its psychological impacts.

The cross-sectional design of the study precludes the possibility of drawing causal inferences, as it is unable to ascertain the direction of the relationships between psychological factors and dizziness. To ascertain causality and the temporal dynamics of these factors, longitudinal studies are required. Although the sample size was relatively large, the generalizability of the findings may be limited due to the combined single-center and online approach of the study and the potential for center-specific biases to influence the results. The use of self-report questionnaires may introduce a degree of bias, and the incorporation of objective assessments could enhance the accuracy of the evaluation. The high prevalence of preoperative dizziness among those with postoperative symptoms suggests that a preexisting condition may affect outcomes. However, the study does not fully examine this interaction. Furthermore, the study does not examine coping mechanisms or interventions that could mitigate the impact of psychological factors on dizziness, underscoring the necessity for future research in this domain.

Nevertheless, it is essential to evaluate the psychometric properties of the DHI, which, despite its pervasive utilization, may exhibit suboptimal properties, as evidenced by extant research (27). Despite the DHI’s limitations, it remains the most commonly utilized tool due to the lack of a more robust alternative in the existing literature. It is recommended that future research focus on validating and enhancing the current measurement tools, as well as investigating effective coping strategies and interventions to more effectively manage the impact of psychological factors on dizziness.

## Conclusion

In conclusion, this study identifies a significant correlation between psychological factors and the severity of dizziness in patients with VSs. It is noteworthy that higher levels of conscientiousness, as well as increased somatization and anxiety, are associated with greater handicap due to dizziness. These findings highlight the necessity of considering psychological factors in the management of postoperative dizziness, with the potential to enhance patient outcomes. Thus, careful assessment of psychological factors especially with regard to premorbid emotional and psychophysiological functioning should be implemented in clinical routine. It would be beneficial for future research to focus on longitudinal studies in order to gain a deeper understanding of the causal relationships between psychological factors and postoperative dizziness in VS patients, as well as to identify effective coping strategies or interventions that could be used to mitigate the impact of these factors on dizziness.

## Data Availability

The data underlying the results presented in the study are available from the first author.

## Ethical approval

All subjects gave their informed consent for inclusion before they participated in the study. The study was conducted in accordance with the Declaration of Helsinki, and the protocol was approved by the Ethics Committee of University Hospital Halle (No. 2020-008).

## Informed Consent Statement

Informed consent was obtained from all subjects involved in the study.

## Data Availability Statement

The data presented in this study are available on request from the corresponding author. The data are not publicly available due to privacy reasons.

## Acknowledgments

We gratefully acknowledge the support provided by Vereinigung Akustikusneurinom e.V.

## References

1. Batuecas-Caletrio A, Santa Cruz-Ruiz S, Muñoz-Herrera A, Perez-Fernandez N. The map of dizziness in vestibular schwannoma. The Laryngoscope. 2015;125(12):2784–9.

2. Carlson ML, Tveiten ØV, Driscoll CL, Neff BA, Shepard NT, Eggers SD, et al. Long-term Dizziness Handicap in Patients with Vestibular Schwannoma: A Multicenter Cross-sectional Study. Otolaryngol Neck Surg. 2014 Dec 1;151(6):1028–37.

3. Fuentealba-Bassaletti C, Neve OM, van Esch BF, Jansen JC, Koot RW, van Benthem PPG, et al. Vestibular Complaints Impact on the Long-Term Quality of Life of Vestibular Schwannoma Patients. Otol Neurotol Off Publ Am Otol Soc Am Neurotol Soc Eur Acad Otol Neurotol. 2023 Feb 1;44(2):161–7.

4. Nilsen KS, Lund-Johansen M, Nordahl SHG, Finnkirk M, Goplen FK. Long-term Effects of Conservative Management of Vestibular Schwannoma on Dizziness, Balance, and Caloric Function. Otolaryngol Neck Surg. 2019 Nov 1;161(5):846–51.

5. Carlson ML, Tveiten ØV, Driscoll CL, Goplen FK, Neff BA, Pollock BE, et al. What drives quality of life in patients with sporadic vestibular schwannoma? The Laryngoscope. 2015;125(7):1697–702.

6. Breivik CN, Nilsen RM, Myrseth E, Finnkirk MK, Lund-Johansen M. Working disability in Norwegian patients with vestibular schwannoma: vertigo predicts future dependence. World Neurosurg. 2013 Dec;80(6):e301–305.

7. Neve OM, Jansen JC, van der Mey AGL, Koot RW, de Ridder M, van Benthem PPG, et al. The impact of vestibular schwannoma and its management on employment. Eur Arch Oto-Rhino-Laryngol Off J Eur Fed Oto-Rhino-Laryngol Soc EUFOS Affil Ger Soc Oto-Rhino-Laryngol - Head Neck Surg. 2022 Jun;279(6):2819–26.

8. Ribeyre L, Spitz E, Frère J, Gauchard G, Parietti-Winkler C. Correlations between postural control and psychological factors in vestibular schwannoma patients. J Vestib Res Equilib Orientat. 2016 Nov 3;26(4):387–94.

9. Saman Y, Mclellan L, Mckenna L, Dutia MB, Obholzer R, Libby G, et al. State Anxiety Subjective Imbalance and Handicap in Vestibular Schwannoma. Front Neurol [Internet]. 2016 Jul 13 [cited 2024 Sep 13];7. Available from: https://www.frontiersin.org/journals/neurology/articles/10.3389/fneur.2016.00101/full

10. Aksoy S, Cekic S. The Relationship Between Vertigo/Dizziness and Somatoform Complaints: A Systematic Review. Indian J Otolaryngol Head Neck Surg. 2024 Feb 1;76(1):1434–46.

11. Kantekin Y, Karaaslan Ö, Dağıstan H, Can İH. Evaluation of anxiety sensitivity, depression, and personality characteristics in chronic subjective dizziness patients. J Surg Med. 2020 Jan 2;4(1):33–7.

12. Trinidade A, Harman P, Stone J, Staab JP, Goebel JA. Assessment of Potential Risk Factors for the Development of Persistent Postural-Perceptual Dizziness: A Case-Control Pilot Study. Front Neurol [Internet]. 2021 Jan 21 [cited 2024 Sep 12];11. Available from: https://www.frontiersin.org/journals/neurology/articles/10.3389/fneur.2020.601883/full

13. Trinidade A, Cabreira V, Goebel JA, Staab JP, Kaski D, Stone J. Predictors of persistent postural-perceptual dizziness (PPPD) and similar forms of chronic dizziness precipitated by peripheral vestibular disorders: a systematic review. J Neurol Neurosurg Psychiatry. 2023 Nov 1;94(11):904–15.

14. Kozak HH, Dündar MA, Uca AU, Uğuz F, Turgut K, Altaş M, et al. Anxiety, Mood, and Personality Disorders in Patients with Benign Paroxysmal Positional Vertigo. Arch Neuropsychiatry. 2018 Mar 19;55(1):49–53.

15. Thomas M, Rampp S, Scheer M, Strauss C, Prell J, Schönfeld R, et al. Premorbid Psychological Factors Associated with Long-Term Postoperative Headache after Microsurgery in Vestibular Schwannoma—A Retrospective Pilot Study. Brain Sci. 2023 Aug 7;13(8):1171.

16. Koos WT, Day JD, Matula C, Levy DI. Neurotopographic considerations in the microsurgical treatment of small acoustic neurinomas. J Neurosurg. 1998;88(3):506–12.

17. Kurre A, Bastiaenen CH, van Gool CJ, Gloor-Juzi T, de Bruin ED, Straumann D. Exploratory factor analysis of the Dizziness Handicap Inventory (German version). BMC Ear Nose Throat Disord. 2010;10:1–10.

18. Robinson K, Gatehouse S, Browning GG. Measuring patient benefit from otorhinolaryngological surgery and therapy. Ann Otol Rhinol Laryngol. 1996;105(6):415–22.

19. McCrae RR, Costa PT. Validation of the five-factor model of personality across instruments and observers. J Pers Soc Psychol. 1987;52(1):81.

20. Muck PM, Hell B, Gosling SD. Ten-Item Personality Inventory--German Version. Eur J Psychol Assess. 2011;

21. Costa Jr PT, McCrae RR. The Revised Neo Personality Inventory (neo-pi-r). Sage Publications, Inc; 2008.

22. Rief W, Hiller W, Heuser J. Screening für somatoforme Störungen (SOMS). Hogrefe Göttingen; 2008.

23. Herrmann-Lingen CBU; Snaith, RP. HADS-D Hospital Anxiety and Depression Scale. Deutsche Version. Bern: Huber; 2002.

24. IBM Corp. IBM SPSS Statistics for Mac. Armonk, NY: IBM Corp; 2021.

25. Benjamini Y, Hochberg Y. Controlling the false discovery rate: a practical and powerful approach to multiple testing. J R Stat Soc Ser B Methodol. 1995;57(1):289–300.

26. Scarff JR, Lippmann S. Treating Psychiatric Symptoms in Persistent Postural Perceptual Dizziness. Innov Clin Neurosci. 2023 Dec 1;20(10–12):49–54.

27. Koppelaar-van Eijsden HM, Schermer TR, Bruintjes TD. Measurement properties of the dizziness handicap inventory: a systematic review. Otol Neurotol. 2022;43(3):e282–97.

